# Impaired sensory evidence accumulation and network function in Lewy body dementia

**DOI:** 10.1101/2021.01.07.21249362

**Authors:** Claire O’Callaghan, Michael Firbank, Alessandro Tomassini, Julia Schumacher, John T O’Brien, John-Paul Taylor

## Abstract

Deficits in attention underpin many of the cognitive and neuropsychiatric features of Lewy body dementia. These attention-related symptoms remain difficult to treat and there are many gaps in our understanding of their neurobiology. An improved understanding of attention-related impairments can be achieved via mathematical modelling approaches, which identify cognitive parameters to provide an intermediate level between observed behavioural data and its underlying neural correlate. Here, we apply this approach to identify the role of impaired sensory evidence accumulation in the attention deficits that characterise Lewy body dementia. In 31 people with Lewy body dementia (including 13 Parkinson’s disease dementia and 18 dementia with Lewy bodies cases), 16 people with Alzheimer’s disease, and 23 healthy controls, we administered an attention task whilst they underwent functional 3T MRI. Using hierarchical Bayesian estimation of a drift diffusion model, we decomposed task performance into drift rate and decision boundary parameters. We tested the hypothesis that the drift rate – a measure of the quality of sensory evidence accumulation – is specifically impaired in Lewy body dementia, compared to Alzheimer’s disease. We further explored whether trial-by-trial variations in the drift rate related to activity within the default and dorsal attention networks, to determine whether altered activity in these networks was associated with slowed drift rates in Lewy body dementia. Our results revealed slower drift rates in the Lewy body dementia compared to the Alzheimer’s disease group, whereas the patient groups were equivalent for their decision boundaries. The patient groups were reduced relative to controls for both parameters. This highlights sensory evidence accumulation deficits as a key feature that distinguishes attention impairments in Lewy body dementia, consistent with impaired ability to efficiently process information from the environment to guide behaviour. We also found that the drift rate was strongly related to activity in the dorsal attention network across all three groups, whereas the Lewy body dementia group showed a divergent relationship relative to the Alzheimer’s disease and control groups for the default network, consistent with altered default network modulation being associated with impaired evidence accumulation. Together, our findings reveal impaired sensory evidence accumulation as a specific marker of attention problems in Lewy body dementia, which may relate to large-scale network abnormalities. By identifying impairments in a specific sub-process of attention, these findings will inform future exploratory and intervention studies that aim to understand and treat attention-related symptoms that are a key feature of Lewy body dementia.

## Introduction

Deficits in attention are a prominent feature of Lewy body dementia – an umbrella term that includes dementia with Lewy bodies and Parkinson’s disease dementia. Core symptoms of these diseases, including a dysexecutive profile, recurrent visual hallucinations and cognitive fluctuations have all been related to attentional impairments (Ballard *et al*., 2002; Mosimann *et al*., 2006; Emre *et al*., 2007; McKeith *et al*., 2017). These pose a difficult constellation of symptoms to treat, and there are still many gaps in our understanding of their underlying neurobiology. A potential way forward is to move away from lumping deficits together under the somewhat nebulous and poorly specified construct of attention dysfunction (Hommel *et al*., 2019; Buzsáki, 2020), and identify more specific processes that are impaired.

One strategy is to apply mathematical modelling approaches that decompose behavioural tasks into their constituent sub-processes (Forstmann *et al*., 2016). Such models aim to provide an intermediate level between observed behavioural data and its underlying neural correlate (Mulder *et al*., 2014). In neurodegenerative conditions, parameters derived from these models have been informative in classifying syndromes (Zhang *et al*., 2016, O’Callaghan *et al*., 2017*a*) and in measuring the effects of treatment interventions (Herz *et al*., 2018; Tomassini *et al*., 2019; O’Callaghan *et al*., 2020). A class of models that can be used to explore sub-processes related to attention are drift diffusion models. In these models, perceptual decision making is conceptualised as the accumulation of noisy sensory evidence over time, until it reaches a threshold where a choice is made (Gold and Shadlen, 2007; Ratcliff and McKoon, 2008; Voss *et al*., 2013). The ability to efficiently select and accumulate sensory information, in order to prioritise and shape interactions with the environment, forms a key component of what is broadly termed “attention” (Feldman and Friston, 2010; Hommel *et al*., 2019).

Processes that contribute to efficient sensory evidence accumulation are affected in Lewy body dementia. Visual dysfunction is common, ranging from deficits in early processes such as contrast and colour discrimination, to impairments in higher-order object perception (Metzler-Baddeley, 2007; Weil *et al*., 2016). Likewise, the co-ordination of brain networks subserving externally vs. internally driven orienting processes are impaired (Shine *et al*., 2014; Gratwicke *et al*., 2015; O’Dowd *et al*., 2019). In Lewy body dementia, there has been a growing focus on the interplay between the default network, which is typically supressed during externally-oriented, demanding tasks, and the dorsal attention system which is engaged during such tasks (Corbetta and Shulman, 2002; Dixon *et al*., 2017; Buckner and DiNicola, 2019). Across the Lewy body disease spectrum, impaired performance on attention-related tasks has been linked to altered co-ordination between the default network and dorsal attention or primary visual networks (Sauer *et al*., 2006; Shine *et al*., 2015; Firbank *et al*., 2016, 2018; Kobeleva *et al*., 2017; Onofrj *et al*., 2019; Walpola *et al*., 2020). Overall, these studies have pointed towards impaired engagement of externally orienting systems, coupled with a reduced ability to modulate the default network. Related to this are findings in Lewy body dementia that show a reduced variability of global brain network efficiency (Schumacher *et al*., 2019*a*) and slowed microstate dynamics (Schumacher *et al*., 2019*b*) – consistent with abnormally rigid brain networks that lack the necessary flexibility to respond to environmental demands, such as sensory evidence accumulation.

Much of the existing work in sensory evidence accumulation has involved single-unit recordings in non-human primates, identifying “accumulator regions” – for example, in the lateral intraparietal area, frontal eye fields and superior colliculus – where activity increases in a ramp-like, or drift, fashion as information is integrated towards a choice (Horwitz and Newsome, 1999; Shadlen and Newsome, 2001; Roitman and Shadlen, 2002; Ding and Gold, 2012). In contrast, fMRI studies have attempted to provide a more holistic, systems-level view of evidence accumulation correlates across the brain (O’Connell *et al*., 2018; Yau *et al*., 2020). These have broadly implicated a frontoparietal network involved in externally oriented attention (Heekeren *et al*., 2004; Ho *et al*., 2009; Liu and Pleskac, 2011; Mulder *et al*., 2014), suggesting that accumulation processes that guide choices rely on the flexible engagement of large scale networks.

Here, we apply a drift diffusion model to a paradigm commonly used to measure attention: the Attention Network Task (Fan *et al*., 2002). We test the hypothesis that the *drift rate*, a measure of the quality of sensory evidence accumulation, is specifically impaired in Lewy body dementia, compared to Alzheimer’s disease. In this way, we contrast two related neurodegenerative conditions that both manifest attentional impairments, to determine whether impaired sensory evidence accumulation may be a specific feature of Lewy body dementia. As participants performed the task whilst undergoing functional MRI, we further explored whether trial-by-trial variations in the drift rate related to activity within the default and dorsal attention networks, to determine whether divergent activity in either, or both, of these networks might be associated with slowed drift rates in Lewy body dementia.

## Methods

### Case selection

The study involved 31 people with Lewy body dementia (13 Parkinson’s disease dementia and 18 dementia with Lewy bodies cases), 16 people with Alzheimer’s disease, and 23 healthy controls. The participants were a subset drawn from our previous study (Firbank *et al*., 2016, 2018), who were selected on the basis of completing a sufficient number of trials in the experimental task (further details below).

The patient groups were prospectively recruited from a population of community-dwelling individuals referred to local neurology and old age psychiatry services, aged 60 years or older with mild to moderate dementia (Mini Mental State Examination (MMSE) score >12). Diagnoses of probable dementia with Lewy bodies and Parkinson’s disease dementia were made using the revised International Consensus Guidelines for dementia with Lewy bodies (McKeith *et al*., 2005) and diagnostic criteria for Parkinson’s disease dementia (Emre *et al*., 2007); probable Alzheimer’s disease was diagnosed based on the National Institute on Aging-Alzheimer’s Association criteria (McKhann *et al*., 2011). Healthy controls were friends or spouses of participants. The study was approved by the local ethics committee and written informed consent was obtained from all participants.

All participants underwent general cognitive assessment using the Cambridge Cognitive Examination (CAMCOG; (Roth *et al*., 1986)) and the MMSE. Presence and severity of extrapyramidal signs was assessed using the motor component of the Unified Parkinson’s disease rating scale (UPDRS-III). Cognitive fluctuations were assessed using the Clinician Assessment of Fluctuation (CAF; (Walker *et al*., 2000*c*)), which measures duration and frequency of fluctuations, and the Mayo Fluctuation Scale (Ferman *et al*., 2004) which includes two dimensions of fluctuations: cognitive-attention and arousal-alertness subscales (Bliwise *et al*., 2014). Patients underwent these general assessments and the fMRI experimental task on their regular medications, with all Lewy body dementia patients in an “on” motor state, typically 1-3 hours after their last dose. Exclusion criteria for all participants included moderate to severe visual impairment, history of alcohol or substance misuse, significant neurological or psychiatric history, moderate to severe cerebral small vessel disease or focal brain lesions on imaging, or the presence of other severe or unstable medical illness. Additional criteria for control participants was an absence of cognitive impairment, based on either self-reported history and/or a score of <80 on the CAMCOG. Demographics and clinical features are reported in Table 1.

**Table 1.** Mean (standard deviation) for demographics and clinical characteristics.

| Demographics & clinical characteristics | Controls | AD | LBD | Group *p* value | Post hoc *p* value |
| --- | --- | --- | --- | --- | --- |
| **N** | 23 | 16 | 31 | - | - |
| **Sex (M:F)** | 16:7 | 13:3 | 25:6 | n.s. | - |
| **Age** | 76.3 (5.4) | 76.4 (8.2) | 75.9 (5.3) | n.s. | - |
| **Education** | 11.4 (1.8) | 10.6 (1.4) | 10.5 (1.5) | n.s. | - |
| **MMSE** | 29.1 (0.8) | 22.7 (3.0) | 24.0 (3.5) | *** | AD vs. Con<sup>***</sup><br>LBD vs. Con<sup>***</sup><br>LBD vs. AD<sup>n.s.</sup> |
| **CAMCOG** | 96.8 (3.5) | 71.6 (12.2) | 77.8 (12.0) | *** | AD vs. Con<sup>***</sup><br>LBD vs. Con<sup>***</sup><br>LBD vs. AD<sup>n.s.</sup> |
| **UPDRS-III** | 1.3 (1.7) | 2.0 (1.6) | 19.3 (7.9) |  | AD vs. Con<sup>n.s.</sup><br>LBD vs. Con<sup>***</sup><br>LBD vs. AD<sup>***</sup> |
| **CAF total** | - | 0.3 (0.8) | 4.4 (3.9) | *** |  |
| **Mayo total** | - | 9.0 (4.6) | 13.3 (6.2) | * |  |
| **Mayo cognitive** | - | 2.1 (1.9) | 2.7 (2.0) | n.s. |  |
| **Mayo arousal** | - | 0.9 (1.0) | 2.3 (1.4) | *** |  |
| **Cholinesterase Inhibitors** | - | 16 (100%) | 27(87%) | - | - |
| **LEDD (mg/day)** | - | - | 683.9 (450.1) | - | - |

n.s. = non significant; \*\*\* = $p < .001$ ; \*\* = $p < .01$ ; \* = $p < .05$ .
Significance tests refer to between group one-way ANOVAs and post hoc Sidak-corrected pairwise t-tests. AD = Alzheimer's disease; LBD = Lewy body disease; MMSE = Mini-Mental State Examination; CAMCOG = Cambridge Cognitive Examination; UPDRS-III = Unified Parkinson's disease rating scale motor component; CAF total = Clinician Assessment of Fluctuation total score; Mayo total = Mayo Fluctuations Scale; Mayo cognitive = Mayo Fluctuation cognitive subscale; Mayo arousal = Mayo Fluctuations arousal subscale; LEDD = levodopa equivalent daily dose.

### Attention Network Task (ANT)

We administered a modified version of the Attention Network Task (ANT; (Fan *et al*., 2002)). The original ANT requires participants to determine the direction of a central arrow flanked by flat lines (neutral) or by arrows pointing in the same (congruent) or different (incongruent) directions. The incongruent condition creates perceptual conflict, which is designed to place greater demand on attentional processes relative to the congruent and neutral conditions. The version used in this study incorporated two levels of perceptual conflict (Firbank *et al*., 2016, 2018). In each trial, participants were shown four arrowheads and had to indicate the direction that the majority were pointing. The four arrowheads were either pointing in the same direction (congruent), or one arrowhead would be pointing in the opposite condition, with its position either at the end of the row (incongruent-EASY) or in the middle of the row (incongruent-HARD, see Fig. 1A). The three conditions provided increasing levels of perceptual conflict and attentional demand. The ANT also contains spatial and warning cues within the three conditions to evaluate alerting and orienting, but these were not analysed in the current study.

**Figure 1.**
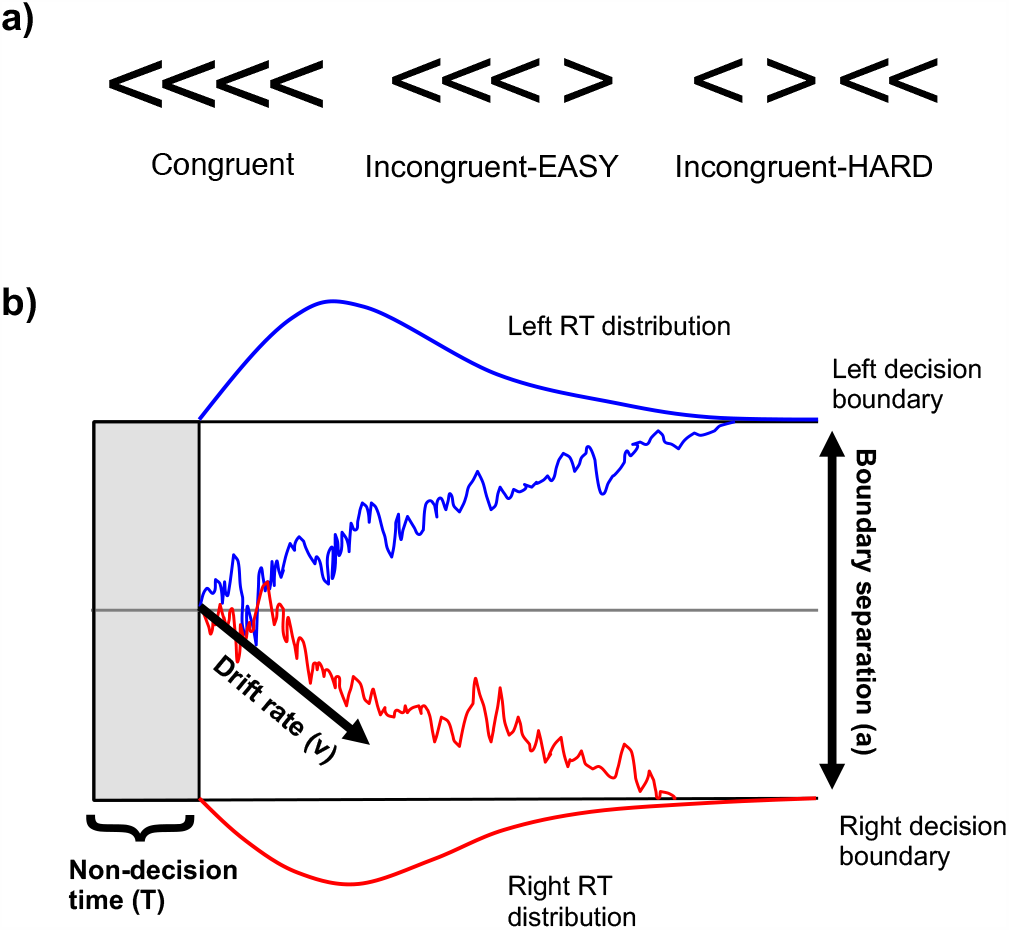
a) Examples of the three conditions in the Attention Network Task; b) Schematic example of drift diffusion model. Two decision boundaries (left and right) are separated by the boundary threshold (*a*). Evidence is nosily accumulated toward a left or right response, with the average evidence accumulation denoted by drift rate (*v*). In this way, the drift rate refers to the average amount of sensory evidence gathered per unit of time (Zhang and Rowe, 2014). The evidence accumulation begins after a period of non-decision time (*T*). Density plots show the distribution of observable reaction times (RT) that are used to calculate the parameters. Adapted from (O’Callaghan *et al*., 2017*a*).

Each run of the task involved 36 trials. All participants included in the current study completed either 5 or 6 runs of the task (a total of 180 or 216 trials). Participants with missed responses on more than 10% of trials were excluded from the current study. These measures ensured that all participants demonstrated adequate task engagement and completed a sufficient number of trials to support drift diffusion modelling.

In the task participants were instructed to respond as quickly and accurately as possible. On each trial a cue was presented for 200 ms, then the target stimulus (i.e., the four arrowheads) was displayed until a response was made or for a maximum of 3000 ms. Response reaction times were recorded up until the appearance of the next cue (which was a minimum of 4300 ms after previous target appearance). Participants indicated their choice by pushing a left or right button. There was a variable delay between the disappearance of the cue and onset of the arrowheads (delays were exponentially distributed at times 700, 770, 850, 960, 1080, 1240, 1430, 1660, 1940, 2300, 2700, 3200 ms, each occurring three times per run in random order). There was also a variable delay between the onset of the arrowheads and the onset of the next cue (at times: 4300, 4500, 4750, 5000, 5350, 5700, 6100, 6400, 6800, 7200, 7700, 8300 ms, each occurring randomly three times per run). The stimuli were back-projected on to a screen at the foot of the scanner and viewed via a mirror positioned at the participants’ eye level. Participants’ best near visual acuity was assessed using Landolt broken rings and fMRI compatible goggles with lenses that ranged from 24.0 to 4.0 diopters (0.5 increment) were used to correct refractive errors. The task was programmed in Matlab using the cogent toolbox (http://www.vislab.ucl.ac.uk/cogent_2000.php).

### Statistical analysis

Analyses were performed in R version 3.3.1 (http://www.r-project.org/). Demographic and clinical variables were analysed using one-way ANOVA and independent samples t-tests. For the ANT, percentage correct and reaction times were analysed in separate mixed effect model repeated measures ANOVAs. The distribution of residuals was checked with Q-Q plots and the Shapiro-Wilk test. Preliminary examination showed that residuals deviated from a normal distribution showing skew and excess leptokurtosis, therefore reaction times (positive skew) were transformed with a log_10_ transformation and percentage correct (negative skewed) were transformed with a square transformation before performing the final analysis (Cardinal and Aitken, 2013). Following significant main effects, pairwise comparisons were made with Sidak corrections. Significant interactions were followed by tests of simple effects.

### Hierarchical drift diffusion model of the ANT

Drift diffusion models (DDMs) can be fitted to rapid, two-choice decision making tasks (Ratcliff and McKoon, 2008; Wagenmakers, 2009; Ratcliff *et al*., 2016). Four main parameters are derived from the DDM: drift rate *v*, decision boundary *a*, decision bias *z* and non-decision time *T*. In a DDM, the decision process is modelled as the accumulation of information, reflected by the drift rate, which continues over time until a decision boundary (i.e., the criterion for how much evidence is required) is reached (Krypotos *et al*., 2015). Decision bias reflects an *a priori* bias toward one of the two responses, and non-decision time comprises those aspects that are not part of the decision making process, including encoding the visual stimulus and executing a motor response (Ratcliff *et al*., 2016). A schematic of the drift diffusion process is illustrated in Fig. 1B).

We applied a hierarchical DDM (hDDM), implemented in python 2.7 using the hDDM toolbox (http://ski.clps.brown.edu/hddm_docs/ (Wiecki *et al*., 2013)) to fit the response and reaction time data from the ANT. The hDDM uses Bayesian estimation to generate posterior distributions of DDM parameters at both the group and individual subject levels. This approach optimises the trade-off between within- and between-subject random effects, accounting for both within-subject variability and group level similarities, as individual parameters are constrained by a group level distribution.

Given the three different levels of perceptual difficulty in the task, we predicted that decision parameters (*v* and *a*) would vary as a function of condition. Given the comparable stimulus encoding and motor requirements across the three conditions, we assumed that non-decision time (*T*) would not vary across conditions. Left and right responses were counterbalanced, so we assumed an unbiased starting point (*z*).

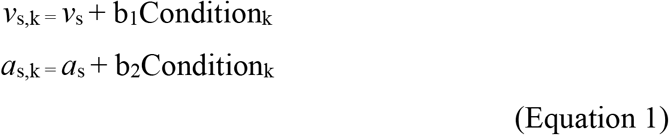

**Equation 1:** Where *v*_s,k_ and *a*_s,k_ are the drift rate and decision boundary of participant *s* on trial *k*. Condition_k_ is the condition (i.e., congruent, incongruent-EASY, incongruent-HARD) on trial *k*, and b_1_ and b_2_ the estimated regression coefficients.

We tested three models: the first allowed drift rate (*v*) to vary by condition, holding decision boundary (*a*) constant; the second allowed *a* to vary across conditions, holding *v* constant; in the third model, *v* and *a* were free to vary across conditions. For each model Markov Chain Monte Carlo simulations generated 95000 samples from the joint posterior parameter distribution, with the first 35000 samples discarded as burn-in, using a thinning factor of 5 with outliers specified at 5%. Convergence was assessed by visual inspection of Markov chains, and with the R-hat Gelman-Rubin statistic where successful convergence is indicated by values <1.1 (Krypotos *et al*., 2015). The best fitting model was determined via the deviance information criterion (DIC) of each model, which evaluates a model’s goodness-of-fit while accounting for complexity (i.e., number of free parameters), with lower DIC values indicating better model fit (Spiegelhalter *et al*., 2002). We ran posterior predictive checks to confirm that the model could reliably reproduce key patterns in the observed data (Wiecki *et al*., 2013). This involved simulating data based on 500 parameter values from the model’s posterior to compare the simulated with the observed data.

Results from the hDDM were analysed using Bayesian hypothesis testing to determine the extent of overlap between the samples drawn from two posterior density distributions. Posterior probabilities can be considered significantly different if < 5% of the distributions overlap (Wiecki *et al*., 2013; Cavanagh *et al*., 2014; Herz *et al*., 2016). The percentage of overlap in the posterior probabilities is denoted by *P* to distinguish it from the classical frequentist *p* values.

### Regression analysis of trial-by-trial network activity on drift rate

The hierarchical drift-diffusion model of the ANT (described in the previous section) identified the drift rate (*v*) as the parameter of interest to differentiate between the Lewy body dementia and Alzheimer’s disease groups. To explore the relationship between fMRI activation and drift rate, we constructed a regression model to determine the relationship between trial-by-trial activation in the dorsal attention and default networks, and the drift rate. This regression model allows estimation of the relationship between trial-by-trial variations in a covariate (e.g., BOLD activation) and the DDM parameters (Cavanagh *et al*., 2011; Wiecki *et al*., 2013; Herz *et al*., 2018). To achieve this, trial-by-trial beta series were extracted from the networks and z-scored (detailed in the following section). As the Lewy body dementia, Alzheimer’s disease and control groups showed similar relationships across the three condition types in the hDDM model described in the previous section, we tested whether trial-by-trial variations in network activity predicted changes in the drift rate irrespective of condition type. As per the previous hDDM model (see Equation 1), we allowed the drift rate and decision boundary to vary across conditions, whilst holding non-decision time constant. Following (Herz *et al*., 2018), for the regression model we estimated posteriors of the regression coefficients for trial-wise regressors at the group level only, in order to account for potential collinearity among model parameters (Frank *et al*., 2015). Separate regression models were run using the beta series values from the default and dorsal attention networks.

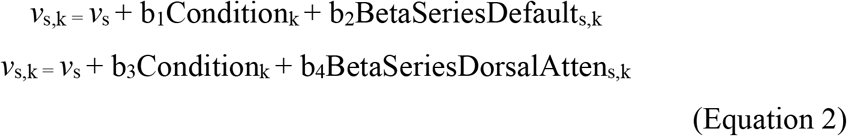

**Equation 2:** Where b_2_BetaSeriesDefault_s,k_ and b_4_BetaSeriesDorsalAtten_s,k_ refer to the extracted beta series from the default and dorsal attention networks, for participant *s* on trial *k*, and *v*_s,k_ is the drift rate.

We evaluated the strength of the relationship between trial-by-trial fluctuations in network activity and drift rate, by determining the extent to which the posterior probability density differed from zero (denoted by *P*, the percentage of the posterior probability distribution different than zero).

### Imaging acquisition

Participants were scanned on a 3 T MRI scanner (Achieva scanner; Philips Medical System, the Netherlands), with body coil transmission and eight channel head coil receiver. A standard whole brain structural scan was acquired (3D MPRAGE, sagittal acquisition, slice thickness 1.0 mm, in plane resolution 1.0 x 1.0 mm; TR = 8.3 ms; TE = 4.6 ms; flip angle = 8°; SENSE factor = 2). fMRI data were collected with a gradient-echo (GE) echo planar imaging (EPI) sequence (TR = 1.92 s; TE = 40 ms; field of view (FOV) = 192 x 192 mm^2^ 64 x 64 matrix size, flip angle 90°, 27 slices, slice thickness 3 mm, slice gap 1 mm) with 156 volumes (5 min).

### fMRI preprocessing and analysis

Imaging analyses were conducted using SPM8 (http://www.fil.ion.ucl.ac.uk/spm/). As described previously in (Firbank *et al*., 2016, 2018), for each participant, the T1 anatomical image was segmented and spatially normalised in SPM using the default parameters. The DARTEL toolbox (Ashburner, 2007) was used to refine the spatial normalisation and create a custom template. fMRI data were first motion corrected by aligning all functional images to the first image for the subject, and subsequently the mean image. Runs with > 3mm or > 3° head motion were excluded. They were then coregistered with the participant’s T1 anatomical image. The spatial normalisation parameters from the T1 image were used to write out the EPI data in standard space with a voxel size of 3 x 3 x 3 mm^3^. The normalised images were then smoothed with an 8 x 8 x 8 mm^3^ FWHM (full width half maximum) Gaussian kernel. A high pass filter of 128 seconds was used, and serial correlations were removed with SPM’s AR(1) model.

We did not perform interpolation or scrubbing of bad image volumes. To investigate possible influences of data quality, we calculated the mean and maximum absolute angular and translational motion between frames (Peraza *et al*., 2015). We also calculated standardised DVARS – the per-image standard deviation of the temporal derivative of the data (Power *et al*., 2012) using Tom Nichols’s script (http://www2.warwick.ac.uk/fac/sci/statistics/staff/academic-research/nichols/scripts/fsl/DVARS.sh) and determined the mean over all volumes in each run. Between the three groups there were no significant differences in mean or maximum xyz motion (mm), mean or maximum xyz angular motion (degree), or mean DVARS (see Supplementary material for details).

The default and dorsal attention networks were identified using the Yeo et al. seven-network parcellation scheme (Yeo *et al*., 2011). To investigate trial-by-trial activation within the networks we extracted beta series. In this approach, each individual trial is entered as a separate regressor in the general linear model (GLM) design matrix, yielding a β estimate of % BOLD signal change for each individual trial (Rissman *et al*., 2004; Cisler *et al*., 2014). To obtain beta series, separate GLMs were performed with the dependent variables being the time course of each network (default and dorsal attention) for each subject, which was calculated as the mean time course for all the voxels in the respective networks. For each run of 36 trials, every target stimulus was modelled as a separate regressor, with six parameters from the motion correction for each functional run included in the design matrix as covariates of no interest. This resulted in individual level beta estimates for each target event, separately for the default and dorsal attention networks. The beta values were z-scored before entering them into the hDDM regression model described above.

### Data availability

Code and data to reproduce the manuscript figures, behavioural analysis and modelling for the Attention Network Task is freely available through the Open Science Framework (https://osf.io/gm8th/).

## Results

### Demographics and clinical characteristics

Demographics and clinical characteristics are detailed in Table 1. The groups were matched for sex (χ^2^ = 1.12, p =0.572), age [F(2, 67) = 0.02, *p* = 0.978] and education level [F(2, 67) = 2.1, *p* = 0.13]. As expected, there were significant group differences in global cognition, on both the MMSE [F(2, 67) = 31.34, *p* < 0.001] and the CAMCOG [F(2, 67) = 35.13, *p* < 0.001], with the Lewy body dementia and Alzheimer’s disease groups performing similar to each other (MMSE: *p*_adjusted_ = 0.160; CAMCOG: *p*_adjusted_ = 0.142), but significantly lower than controls (MMSE: *p*_adjusted_ = < 0.001; CAMCOG: *p*_adjusted_ values < 0.001). Also as expected the severity of motor symptoms differed across the groups [UPDRS-III: F(2, 67) = 89.17, *p* < 0.001], with the Lewy body dementia group showing significantly higher motor severity than both controls and Alzheimer’s disease (*p*_adjusted_ values < 0.001), who did not differ from each other (*p*_adjusted_ = 0.973). The Lewy body dementia group had significantly more fluctuations compared to the Alzheimer’s disease group [CAF: t(33.27) = −5.47, *p* < 0.001; Mayo total: t(39.48) = −2.69, *p* = 0.01]. Within the Mayo assessment, the dementia groups did not differ on the cognitive-attention subscale [Mayo cognitive: t(31.53) = −0.99, *p* = 0.329], but the Lewy body group scored higher on the arousal-alertness subscale [Mayo arousal: t(40.58) = −4.12, *p* < 0.001].

### Attention Network Task behavioural results

#### Percentage correct

Trials where no response was made were excluded from the analysis. The amount of no response trials excluded did not differ significantly across the groups [F(2, 35) = 1.97, *p* = 0.154, see Supplementary Fig. 1].

Fig. 2 (left panel) shows the percentage of correct responses across the groups. Percentage correct scores were subjected to a square transformation to reduce skew and leptokurtosis. Results of the mixed model ANOVA showed a main effect of group [F(2, 67) = 13.76, *p* < 0.001]; post hoc comparisons indicated that the Lewy body dementia group had significantly fewer correct responses compared to controls (*p*_adjusted_ < 0.001) and Alzheimer’s disease (*p*_adjusted_ < 0.05), and the Alzheimer’s disease group had significantly fewer correct responses than controls (*p*_adjusted_ < 0.05).

**Figure 2.**
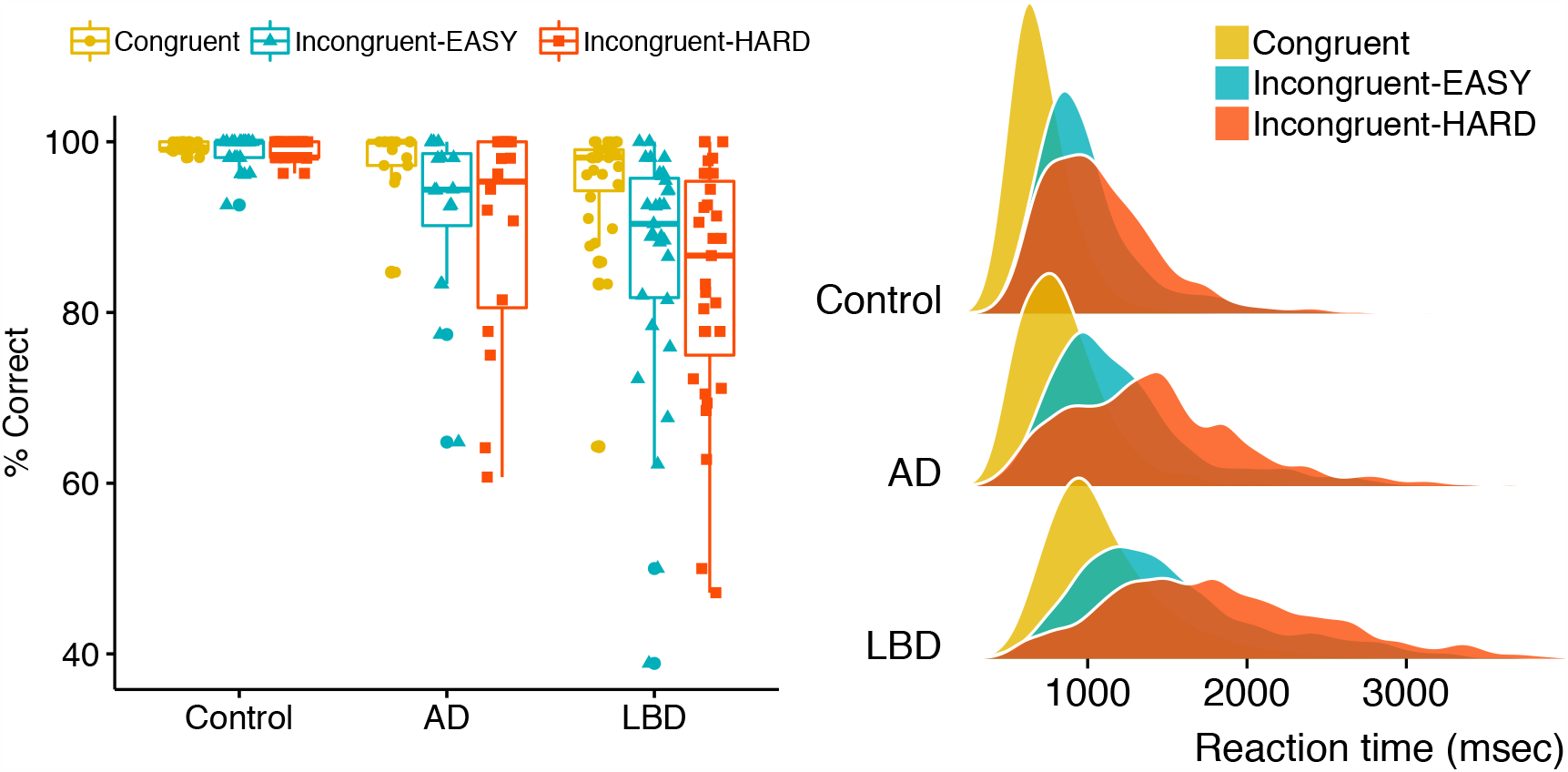
Attention Network Task behavioural results. Left panel: Percentage of correct responses across the three conditions; Right panel: Distributions of reaction times across the three conditions.

There was also a main effect of condition [F(2, 134) = 24.49, *p* < 0.001], with significantly more correct responses achieved in the congruent condition, relative to incongruent-EASY (*p*_adjusted_ < 0.05) and incongruent-HARD (*p*_adjusted_ < 0.001), correct responses did not differ between the incongruent conditions (*p*_adjusted_ = 0.695).

Finally, there was a significant interaction [F(4, 134) = 6.30, *p* < 0.001], which reflected that the patient groups performed worse with respect to controls in the more difficult conditions. Tests of simple effects revealed a significant group difference at each condition: congruent [simple effect, F(2,67) = 4.77, p < 0.05], incongruent-EASY [simple effect, F(2,67) = 10.66, p < 0.001] and incongruent-HARD [simple effect, F(2,67) = 14.11, p < 0.001]. Pairwise comparisons showed that for the congruent condition, the Lewy body dementia group had significantly fewer correct responses than controls [*p*_adjusted_ < 0.05; Lewy body dementia vs. Alzheimer’s disease: *p*_adjusted_ = 0.221; Alzheimer’s disease vs. controls *p*_adjusted_ = 0.773]; for the incongruent-EASY condition the Lewy body dementia group had significantly fewer correct responses than controls [*p*_adjusted_ < 0.001; Lewy body dementia vs. Alzheimer’s disease: *p*_adjusted_ = 0.198; Alzheimer’s disease vs. controls *p*_adjusted_ = 0.099]; and for the incongruent-HARD condition the Lewy body dementia group had significantly fewer correct responses than controls [*p*_adjusted_ < 0.001; Lewy body dementia vs. Alzheimer’s disease: *p*_adjusted_ = 0.210] as did the Alzheimer’s disease group [Alzheimer’s disease vs. controls *p*_adjusted_ < 0.05].

#### Reaction time

Fig. 2 (right panel) shows the distribution of reaction times across the groups. Reaction times were not normally distributed, with positive skew and slight leptokurtosis, but satisfied normality tests following a log_10_ transformation.

Results of the mixed model ANOVA showed a main effect of group [F(2, 67) = 28.50, *p* < 0.001]; post hoc comparisons indicated that the Lewy body dementia group was significantly slower than controls (*p*_adjusted_ < 0.001) and Alzheimer’s disease (*p*_adjusted_ < 0.001), and the Alzheimer’s disease group was significantly slower than controls (*p*_adjusted_ < 0.001). There was also a main effect of condition [F(2, 134) = 384.81, *p* < 0.001], post hoc comparisons showed that responses in both incongruent conditions were significantly slower than in the congruent condition (*p*_adjusted_ values < 0.001); also, responses were slower in the HARD vs. EASY incongruent conditions (*p*_adjusted <_ 0.05). The group x condition interaction was not significant [F(4, 134) = 2.28, *p* = 0.06].

### Hierarchical drift-diffusion model fit

All three models showed good convergence, based on visually inspected chains and all R-hat values < 1.1. Based on the DIC values, the best fitting model was model three where both *v* and *a* varied by condition (DIC model 1: 17089.50; DIC model 2: 17877.63; DIC model 3: 7236.97). Posterior predictive checks revealed agreement between the simulated and observed data (see Supplementary Fig. 2 for observed data plotted against predicted model data).

#### Analysis of hierarchical drift-diffusion model parameters

Fig. 3 shows group comparisons of posterior probability density plots for the drift rate *v* (top panel) and decision boundaries *a* (bottom panel) across each condition.

**Figure 3.**
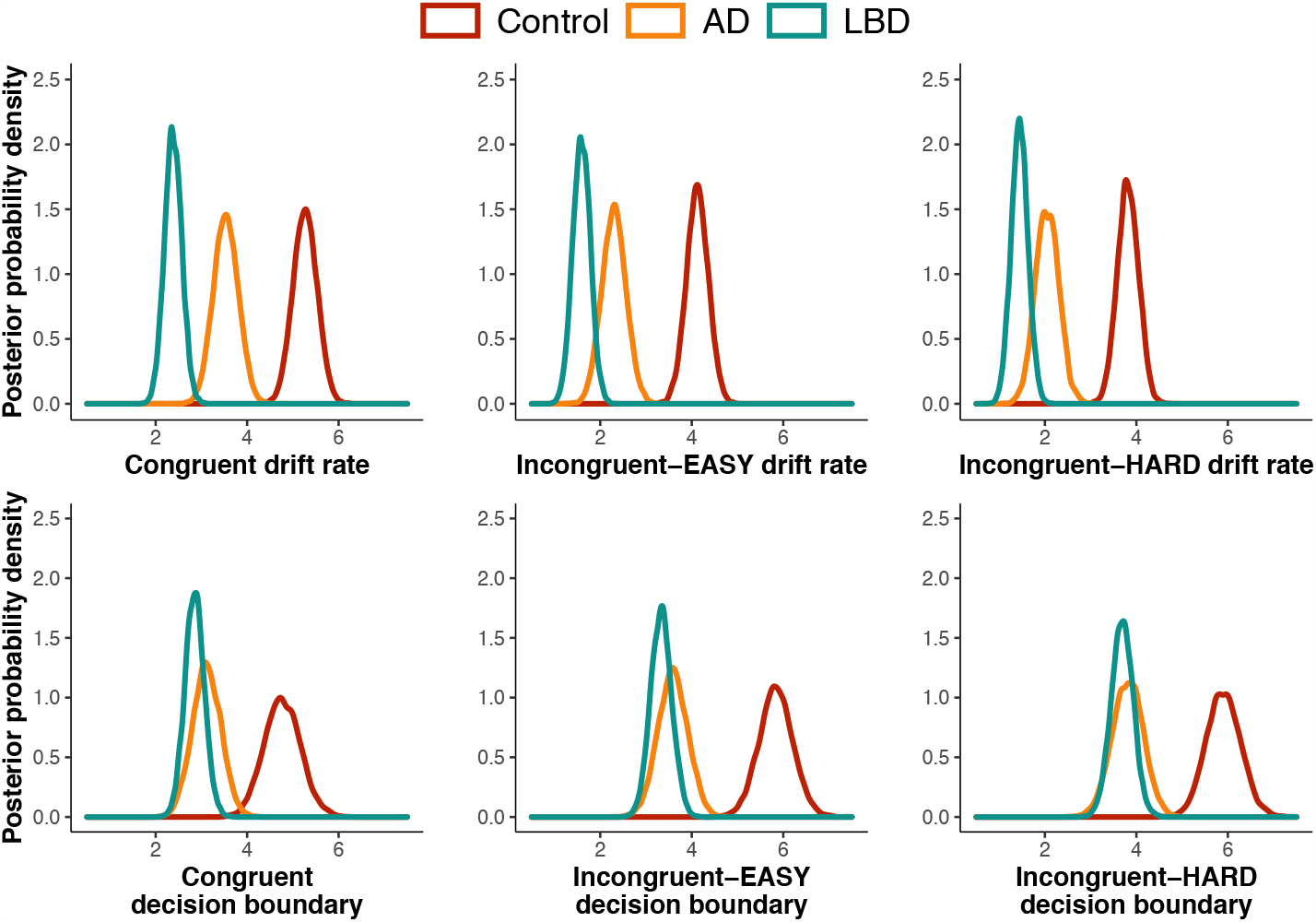
Drift rates and decision boundaries. Group comparisons of the posterior probability density plots for drift rates *v* (top panel) and decision boundaries *a* (bottom panel), in each task condition. Peaks of distributions reflect the most likely value of the parameter.

The disease groups differed consistently in their drift rates, with the Lewy body dementia group showing significantly lower drift rates than Alzheimer’s disease in each condition (congruent: *P* = 0.02%; incongruent-EASY: *P* = 1.35%; incongruent-HARD: *P* = 2.80%). Both the Lewy body dementia and Alzheimer’s disease groups had significantly lower drift rates relative to controls (congruent: *P* = 0.00%; incongruent-EASY: *P* = 0.00%; incongruent-HARD: *P* = 0.00%).

In contrast, the Lewy body dementia and Alzheimer’s disease groups showed considerable overlap in their decision boundaries across all of the conditions (congruent: *P* = 23.49%; incongruent-EASY: *P* = 25.77%; incongruent-HARD: *P* = 38.22%). Both groups had significantly smaller decision boundaries compared to controls (values for Lewy body dementia vs. controls were congruent: *P* = 0.00%; incongruent-EASY: *P* = 0.00%; incongruent-HARD: *P* = 0.00%; values for Alzheimer’s disease vs. controls were congruent: *P* = 0.06%; incongruent-EASY: *P* = 0.00%; incongruent-HARD: *P* = 0.00%).

For the non-decision time (*T*), shown in Fig. 4, all groups differed significantly with the Lewy body dementia group showing the longest non-decision time, followed by the Alzheimer’s disease group, then controls (Lewy body dementia vs. Alzheimer’s disease: *P* = 0.50%; Lewy body dementia vs. controls: *P* = 0.00%; AD vs. controls: *P* = 0.08%).

**Figure 4.**
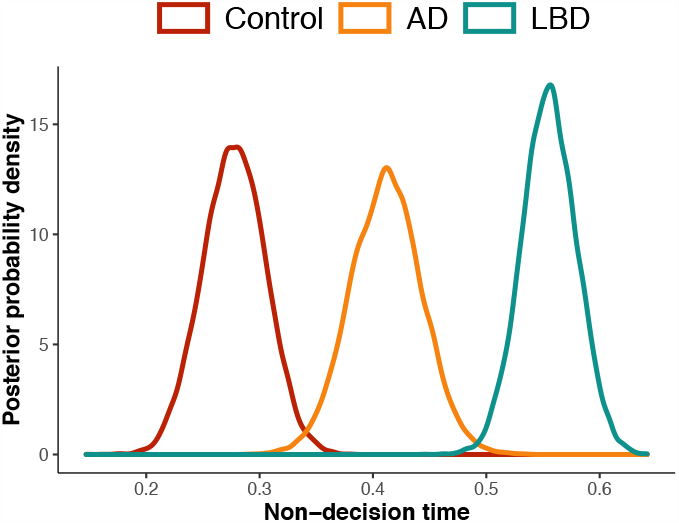
Non-decision time. Group comparison of the posterior probability density plots for non-decision time (*T*), combined across task conditions. Peaks of distributions reflect the most likely value of the parameter.

### Correlations with clinical measures of cognitive fluctuation

Spearman’s correlations were conducted between fluctuation scores (i.e., CAF total, Mayo total, Mayo cognitive-attention and arousal-alertness subscales) in the Lewy body dementia group and mean drift rates across the three task conditions. We did not observe significant correlations in any of these measures (*p* values > 0.95).

### Relationship between drift rate and trial-by-trial fluctuations in network activity

The hDDM identified the drift rate (*v)* as the decision parameter that distinguished Lewy body dementia patients from both the Alzheimer’s disease and control group. We therefore entered beta series derived from the default and dorsal attention networks into an hDDM regression model, in order to determine how trial-by-trial fluctuations in BOLD activity related to changes in the drift rate.

As shown in Fig. 5, for all groups activity in the dorsal attention network was strongly (positively) associated with drift rates, consistent with greater activity in the dorsal attention network being associated with a higher drift rate. This is reflected by the percentage of the posterior probability for all parameters being considerably greater than zero, *P* refers to the percentage of the distribution that is greater (or less than) zero (controls: *P* = 99.92 %; AD: *P* = 94.02 %; Lewy body dementia: *P* = 99.81 %).

**Figure 5.**
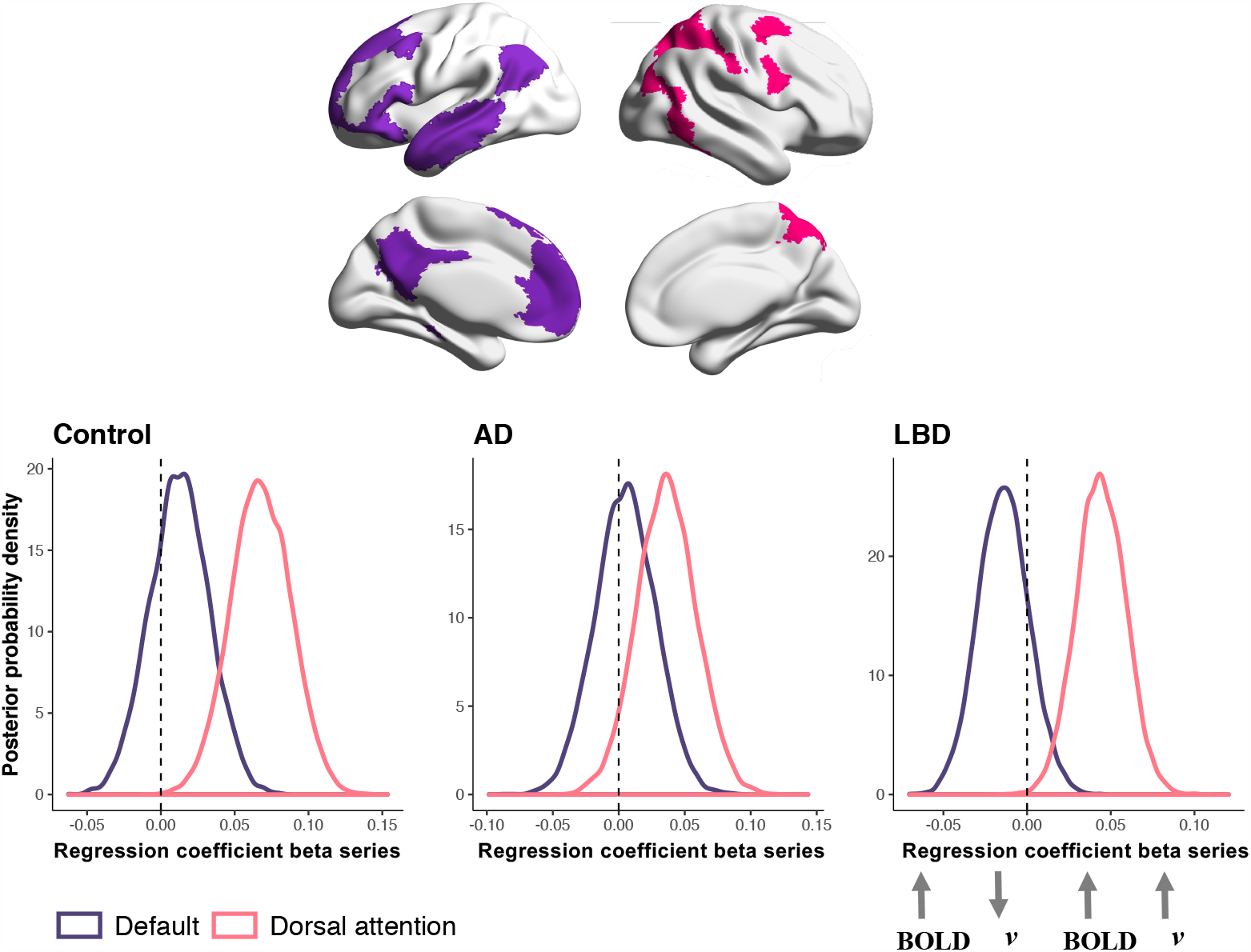
Relationship between drift rate and activity in the default and dorsal attention networks. Top panel shows the default (purple) and dorsal attention (pink) network maps taken from the Yeo et al parcellation (Yeo *et al*., 2011). Bottom panel shows the posterior probability density plots of the drift rates estimated from the hDDM regression model (y-axis) and how they varied as a function of activity in the default and dorsal attention networks (x-axis). Strength of the relationship reflected by the amount of distribution being to the left or right of zero. Peaks of distributions reflect the most likely value of the parameter.

For the default network, as shown in Fig. 5, the majority of the posterior probability densities for controls was to the right of zero, consistent with greater default network activity being positively associated with the drift rate (*P* = 70.96 %); Alzheimer’s disease also showed a rightward shift, but to a lesser extent (*P* = 59.78 %). In contrast, for Lewy body dementia the majority of the posterior probability density was left of zero (*P* = 80.62 %), consistent with greater activity in the default network being associated with a slower drift rate. Although none of these reached the commonly used metric of “significance” designated when > 95% of the posterior density exceeds zero (Cavanagh *et al*., 2014).

## Discussion

We show that the drift rate, a measure of the quality of sensory evidence accumulation, is impaired in Lewy body dementia relative to Alzheimer’s disease. By decomposing decisions in an attentional task into the sub-processes of evidence accumulation (drift rate) and evidence criteria (decision boundary), our results revealed slower drift rates in the Lewy body dementia group, whereas the patient groups were equivalent for their decision boundaries. This highlights that deficits in sensory evidence accumulation may be a key process that distinguishes attention impairments in Lewy body dementia. We further show that the drift rate was strongly related to activity in the dorsal attention network across all three groups, whereas the Lewy body dementia group showed a divergent relationship for the default network, consistent with altered default network modulation being associated with impaired evidence accumulation.

The drift rate measures how efficiently information is accumulated and reflects the quality of evidence that enters the decision making process. In this way, it speaks to the quality of evidence extracted from a stimulus and the level of noise in the evidence accumulation process, capturing whether decisions are fast and accurate, or slow and error prone (Krypotos *et al*., 2015). Both of the disease groups showed a reduced drift rate relative to controls, with the Lewy body dementia group further reduced compared to the Alzheimer’s disease group. These neurodegenerative conditions both manifest impaired attention as part of a dysexecutive cognitive profile (Metzler-Baddeley, 2007), however the striking attention-related symptoms of visual hallucinations and cognitive fluctuations are more common in Lewy body dementia (Walker *et al*., 2000*a*; Ballard *et al*., 2001; Tiraboschi *et al*., 2006). Prominent impairments in sensory evidence accumulation may be a key feature underpinning the symptoms that distinguish Lewy body dementia from related neurodegenerative diseases.

In contrast to the drift rate, both the Lewy body dementia and Alzheimer’s disease groups showed similarly reduced decision boundaries relative to controls. This parameter is considered a measure of response caution, indicating the amount of evidence that needs to be accumulated before a decision is reached. Wider boundaries promote slow but accurate decisions, whereas narrow boundaries are associated with fast but error prone decisions (Starns and Ratcliff, 2010). The narrower boundaries in our patient groups are in keeping with the increased errors they made on the task, relative to controls. Previous work in Parkinson’s disease patients (without dementia) showed they were able to flexibly adjust decision boundaries in response to task demands (Cavanagh *et al*., 2011; Herz *et al*., 2016), with evidence that they set wider decision boundaries compared to age-matched controls (O’Callaghan *et al*., 2017*a*). This is consistent with an amplification of what is seen in healthy ageing: older adults tend to have wider decision boundaries, enabling a conservative decision criteria, which acts as a compensatory strategy to guard against errors in speed-accuracy trade-off tasks (Spaniol *et al*., 2006; Ratcliff *et al*., 2007, 2010). That we show reduced decision boundaries in both dementia patient groups suggests that this ability to compensate via adjusting decision boundaries may decline with advancing cognitive impairment.

With respect to the non-decision time, the Lewy body dementia group had much longer non-decision times compared to the Alzheimer’s group, who were increased relative to controls. This is unsurprising given that non-decision time comprises those components unrelated to the decision itself, including stimulus processing and motor execution – both of which may be particularly impaired in Lewy body dementia, consistent with their characteristic visual processing deficits and extrapyramidal features.

Taken together, our analysis decomposes the raw behavioural results from the Attention Network Task – which indicated slowed reaction times and increased errors in Lewy body dementia – to reveal that their impaired perceptual decision-making was underpinned by alterations in sensory evidence accumulation. As opposed to a broadly defined attentional impairment or cognitive slowing, this result points towards a specific deficit in efficiently processing information from the environment to guide behaviour.

Consistent with the notion that sensory evidence accumulation deficits might contribute to some of the distinguishing features of Lewy body disease, impaired sensory evidence accumulation during perceptual decision making was found in Parkinson’s disease patients (without dementia) who experience visual hallucinations, relative to those who don’t hallucinate (O’Callaghan *et al*., 2017*a*). This is in keeping with a theoretical framework for visual hallucinations where poor quality, or imprecise, sensory information renders the perceptual process vulnerable to excessive influence from prior beliefs (Friston, 2005; Shine *et al*., 2014, O’Callaghan *et al*., 2017*b*; Zarkali *et al*., 2019). Speculatively, the cognitive fluctuations characteristic of Lewy body dementia, which can occur over days or hours, but also over seconds to minutes (Walker *et al*., 2000*b*), may produce transient reductions in sensory evidence accumulation ability. These periods are consistent with a brain state that is less responsive to the environment, potentially reflecting a temporal mismatch between sensory processing demands and the speed and efficiency of intrinsic information processing (Firbank *et al*., 2018, Schumacher *et al*., 2019*b*). Such periods of reduced engagement with the environment may be synonymous with reduced sensory evidence accumulation, and future work could explore this parameter as a possible marker of cognitive fluctuations. We have shown clear differences for the drift rates in a fluctuating dementia group (Lewy body dementia) versus a non-fluctuating group (Alzheimer’s disease). However, the extent of drift rate slowing did not correlate with clinical measures of fluctuation severity. This suggests that fluctuations on the trial-by-trial timescale that are captured by the drift rate may not be well represented by clinical scales. An important avenue for future work is to establish concordance between different measurement tools across the varying timescales of fluctuations (Matar *et al*., 2020). In this respect, the hierarchical drift diffusion model offers a unique possibility to capture objective moment-by-moment fluctuations, in contrast to other neuropsychological tools that summarise performance across trials.

Our imaging analysis showed that the drift rate was strongly related to trial-by-trial fluctuations in BOLD activity in the dorsal attention network for all three groups. That is, increased BOLD activation within the dorsal attention network was associated with a faster drift rate – and this relationship was preserved in both the Alzheimer’s disease and Lewy body dementia groups. Anatomically, the dorsal attention network overlaps with frontal eye fields and the intraparietal sulcus, which is a human homologue of the monkey lateral intraparietal region (Grefkes and Fink, 2005), and therefore the network overlaps with regions previously identified as showing ramping neuronal activity during evidence accumulation (Shadlen and Newsome, 2001; Roitman and Shadlen, 2002; Ding and Gold, 2012).

The positive relationship between drift rate and BOLD activity in the dorsal attention network is in keeping with single-unit studies where firing rates increase as information is integrated towards a decision. However, given the comparative slowness of the BOLD signal, and the pooled neuronal populations it represents, how evidence accumulation measured by single-unit recordings should be expressed in fMRI signals remains a matter of debate (Mulder *et al*., 2014; Hanks and Summerfield, 2017; O’Connell *et al*., 2018). Indeed, while some studies have related higher BOLD responses to increasing evidence accumulation as we found (Heekeren *et al*., 2004; Philiastides and Sajda, 2007), others have shown an inverse relationship between BOLD and the drift rate (Ho *et al*., 2009; Liu and Pleskac, 2011). The inverse relationship can be interpreted as BOLD reflecting the pooled activity of neurons, such that the highest aggregate activity will be observed when sensory evidence is weak/noisy and the build-up of firing, although shallower, will be prolonged with slower drift rates, resulting in increased overall BOLD on those trials (Mulder *et al*., 2014; Hanks and Summerfield, 2017). Whilst our finding does not disambiguate this debate, it establishes a positive relationship between dorsal attention network activity and drift rate during perceptual decision making, in both healthy elderly and neurodegenerative disease cohorts.

With respect to the default network, the groups showed divergent responses. The control and Alzheimer’s disease groups showed a weak, positive relationship between the drift rate and BOLD activity within the default network. In contrast, the Lewy body dementia group showed a stronger, negative relationship, consistent with greater default network activity being associated with a slower drift rate. Whilst these results did not reach the commonly used “significance” criterion, the difference in directionality suggests a tentative mechanistic interpretation: that increased default network activity during perceptual decision making is associated with impaired evidence accumulation in Lewy body dementia. This finding is consistent with other work showing altered default network modulation during attention-related tasks across the Lewy body disease spectrum (Sauer *et al*., 2006; Shine *et al*., 2015; Firbank *et al*., 2016, 2018), and suggests that impaired sensory evidence accumulation may be a specific behavioural correlate of default network dysfunction in Lewy body dementia. In Lewy body disease, symptoms associated with attentional impairments, including dysexecutive problems, fluctuations and visual hallucinations, are increasingly conceptualised as disturbances in large-scale brain networks, driven by local pathological changes and dysfunctional modulation from ascending neurotransmitter systems (Shine *et al*., 2014; Gratwicke *et al*., 2015; O’Dowd *et al*., 2019; Matar *et al*., 2020). Improved understanding of large-scale network dysfunction in these symptoms may continue to shape future treatment options, given the potential to modulate networks via drugs or brain stimulation (Gratwicke *et al*., 2015). Our findings suggest that impaired sensory evidence accumulation is a specific marker of attention problems in Lewy body dementia, which may relate to large-scale network engagement. Going forward, specific behavioural parameters, such as the drift rate, may be important to exploit in explanatory and treatment studies of attention-related impairments in Lewy body dementia.

## Data Availability

Data availability Code and data to reproduce the manuscript figures, behavioural analysis and modelling for the Attention Network Task is freely available through the Open Science Framework (https://osf.io/gm8th/).

## Acknowledgments

We thank Alison Killen for the initial data collection.

## Funding

CO was supported by a Neil Hamilton Fairley Fellowship from the Australian National Health and Medical Research Council (GNT1091310). AT was supported by a UK Medical Research Council Intramural Programme Grant (SUAG/051). JOB is supported by the NIHR Cambridge Biomedical Research Centre and the Cambridge Centre for Parkinson’s Plus. The research was supported by a Wellcome Trust Intermediate Clinical Fellowship (WT088441MA) to J-PT, Northumberland Tyne and Wear NHS Foundation Trust, by National Institute for Health Research (NIHR) Newcastle Biomedical Research Centre (BRC) based at Newcastle upon Tyne Hospitals NHS Foundation Trust and Newcastle University, and by Alzheimer’s Research UK.

## Notes

### Competing Interest Statement

The authors have declared no competing interest.

### Author Declarations

NHS Health Research Authority: North East - Newcastle and North Tyneside

## References

Ashburner J. A fast diffeomorphic image registration algorithm. NeuroImage 2007; 38: 95–113.

Ballard C, O’Brien J, Gray A, Cormack F, Ayre G, Rowan E, et al. Attention and Fluctuating Attention in Patients With Dementia With Lewy Bodies and Alzheimer Disease. Arch Neurol 2001; 58: 977.

Ballard CG, Aarsland D, McKeith I, O’Brien J, Gray A, Cormack F, et al. Fluctuations in attention: PD dementia vs DLB with parkinsonism. Neurology 2002; 59: 1714–20.

Bliwise DL, Scullin MK, Trotti LM. Fluctuations in cognition and alertness vary independently in dementia with Lewy bodies. Movement Disorders 2014; 29: 83–9.

Buckner RL, DiNicola LM. The brain’s default network: updated anatomy, physiology and evolving insights. Nat Rev Neurosci 2019: 1–16.

Buzsáki G. The Brain–Cognitive Behavior Problem: A Retrospective. eNeuro 2020; 7: ENEURO.0069-20.2020.

Cardinal RN, Aitken MRF. ANOVA for the behavioral sciences researcher. 2013

Cavanagh JF, Wiecki TV, Cohen MX, Figueroa CM, Samanta J, Sherman SJ, et al. Subthalamic nucleus stimulation reverses mediofrontal influence over decision threshold. Nature Neuroscience 2011; 14: 1462–7.

Cavanagh JF, Wiecki TV, Kochar A, Frank MJ. Eye tracking and pupillometry are indicators of dissociable latent decision processes. Journal of Experimental Psychology: General 2014; 143: 1476–88.

Cisler JM, Bush K, Steele JS. A comparison of statistical methods for detecting context-modulated functional connectivity in fMRI. NeuroImage 2014; 84: 1042–52.

Corbetta M, Shulman GL. Control of goal-directed and stimulus-driven attention in the brain. Nat Rev Neurosci 2002; 3: 201–15.

Ding L, Gold JI. Neural Correlates of Perceptual Decision Making before, during, and after Decision Commitment in Monkey Frontal Eye Field. Cereb Cortex 2012; 22: 1052–67.

Dixon ML, Andrews-Hanna JR, Spreng RN, Irving ZC, Mills C, Girn M, et al. Interactions between the default network and dorsal attention network vary across default subsystems, time, and cognitive states. NeuroImage 2017; 147: 632–49.

Emre M, Aarsland D, Brown R, Burn DJ, Duyckaerts C, Mizuno Y, et al. Clinical diagnostic criteria for dementia associated with Parkinson’s disease. Movement Disorders 2007; 22: 1689–707.

Fan J, McCandliss BD, Sommer T, Raz A, Posner MI. Testing the Efficiency and Independence of Attentional Networks. Journal of Cognitive Neuroscience 2002; 14: 340–7.

Feldman H, Friston KJ. Attention, Uncertainty, and Free-Energy [Internet]. Front Hum Neurosci 2010; 4[cited 2020 Oct 16] Available from: http://journal.frontiersin.org/article/10.3389/fnhum.2010.00215/abstract

Ferman TJ, Smith GE, Boeve BF, Ivnik RJ, Petersen RC, Knopman D, et al. DLB fluctuations: specific features that reliably differentiate DLB from AD and normal aging. Neurology 2004; 62: 181–7.

Firbank M, Kobeleva X, Cherry G, Killen A, Gallagher P, Burn DJ, et al. Neural correlates of attention-executive dysfunction in lewy body dementia and Alzheimer’s disease. Hum Brain Mapp 2016; 37: 1254–70.

Firbank MJ, O’Brien JT, Taylor J-P. Long reaction times are associated with delayed brain activity in lewy body dementia. Hum Brain Mapp 2018; 39: 633–43.

Forstmann BU, Ratcliff R, Wagenmakers E-J. Sequential Sampling Models in Cognitive Neuroscience: Advantages, Applications, and Extensions. Annual Review of Psychology 2016; 67: 641–66.

Frank MJ, Gagne C, Nyhus E, Masters S, Wiecki TV, Cavanagh JF, et al. fMRI and EEG Predictors of Dynamic Decision Parameters during Human Reinforcement Learning. J Neurosci 2015; 35: 485–94.

Friston KJ. Hallucinations and perceptual inference. Behavioral and Brain Sciences 2005; 28: 764–6.

Gold JI, Shadlen MN. The Neural Basis of Decision Making. Annual Review of Neuroscience 2007; 30: 535–74.

Gratwicke J, Jahanshahi M, Foltynie T. Parkinson’s disease dementia: a neural networks perspective. Brain 2015; 138: 1454–76.

Grefkes C, Fink GR. The functional organization of the intraparietal sulcus in humans and monkeys. Journal of Anatomy 2005; 207: 3–17.

Hanks TD, Summerfield C. Perceptual Decision Making in Rodents, Monkeys, and Humans. Neuron 2017; 93: 15–31.

Heekeren HR, Marrett S, Bandettini PA, Ungerleider LG. A general mechanism for perceptual decision-making in the human brain. Nature 2004; 431: 859–62.

Herz DM, Little S, Pedrosa DJ, Tinkhauser G, Cheeran B, Foltynie T, et al. Mechanisms Underlying Decision-Making as Revealed by Deep-Brain Stimulation in Patients with Parkinson’s Disease. CURBIO 2018; 28: 1169-1178.e6.

Herz DM, Zavala BA, Bogacz R, Brown P. Neural Correlates of Decision Thresholds in the Human Subthalamic Nucleus. Current Biology 2016; 26: 916–20.

Ho TC, Brown S, Serences JT. Domain General Mechanisms of Perceptual Decision Making in Human Cortex. Journal of Neuroscience 2009; 29: 8675–87.

Hommel B, Chapman CS, Cisek P, Neyedli HF, Song J-H, Welsh TN. No one knows what attention is. Atten Percept Psychophys 2019; 81: 2288–303.

Horwitz GD, Newsome WT. Separate Signals for Target Selection and Movement Specification in the Superior Colliculus. Science 1999; 284: 1158–61.

Kobeleva X, Firbank M, Peraza L, Gallagher P, Thomas A, Burn DJ, et al. Divergent functional connectivity during attentional processing in Lewy body dementia and Alzheimer’s disease. CORTEX 2017; 92: 8–18.

Krypotos A-M, Beckers T, Kindt M, Wagenmakers E-J. A Bayesian hierarchical diffusion model decomposition of performance in Approach-Avoidance Tasks. Cognition and Emotion 2015; 29: 1424–44.

Liu T, Pleskac TJ. Neural correlates of evidence accumulation in a perceptual decision task. Journal of Neurophysiology 2011; 106: 2383–98.

Matar E, Shine JM, Halliday GM, Lewis SJG. Cognitive fluctuations in Lewy body dementia: towards a pathophysiological framework. Brain 2020; 143: 31–46.

McKeith IG, Boeve BF, Dickson DW, Halliday G, Taylor J-P, Weintraub D, et al. Diagnosis and management of dementia with Lewy bodies: Fourth consensus report of the DLB Consortium. Neurology 2017; 89: 88–100.

McKeith IG, Dickson DW, Lowe J, Emre M, O’Brien JT, Feldman H, et al. Diagnosis and management of dementia with Lewy bodies: third report of the DLB Consortium. Neurology 2005; 65: 1863–72.

McKhann GM, Knopman DS, Chertkow H, Hyman BT, Jack Jr. CR, Kawas CH, et al. The diagnosis of dementia due to Alzheimer’s disease: Recommendations from the National Institute on Aging-Alzheimer’s Association workgroups on diagnostic guidelines for Alzheimer’s disease. Alzheimer’s & Dementia 2011; 7: 263–9.

Metzler-Baddeley C. A Review of Cognitive Impairments in Dementia with Lewy Bodies Relative to Alzheimer’s Disease and Parkinson’s Disease with Dementia. Cortex 2007; 43: 583–600.

Mosimann UP, Rowan EN, Partington CE, Collerton D, Littlewood E, O’Brien JT, et al. Characteristics of Visual Hallucinations in Parkinson Disease Dementia and Dementia With Lewy Bodies. The American Journal of Geriatric Psychiatry 2006; 14: 153–60.

Mulder MJ, van Maanen L, Forstmann BU. Perceptual decision neurosciences – A model-based review. Neuroscience 2014; 277: 872–84.

O’Callaghan C, Hall JM, Tomassini A, Muller AJ, Walpola IC, Moustafa AA, et al. Visual Hallucinations Are Characterized by Impaired Sensory Evidence Accumulation: Insights From Hierarchical Drift Diffusion Modeling in Parkinson’s Disease. Biological Psychiatry: Cognitive Neuroscience and Neuroimaging 2017a; 2: 680–8.

O’Callaghan C, Hezemans FH, Ye R, Rua C, Jones PS, Murley AG, et al. Locus coeruleus integrity and the effect of atomoxetine on response inhibition in Parkinson’s disease [Internet]. 2020 [cited 2020 Dec 27] Available from: http://medrxiv.org/lookup/doi/10.1101/2020.09.03.20176800

O’Callaghan C, Kveraga K, Shine JM, Adams RB, Bar M. Predictions penetrate perception: Converging insights from brain, behaviour and disorder. Consciousness and Cognition 2017b; 47: 63–74.

O’Connell RG, Shadlen MN, Wong-Lin K, Kelly SP. Bridging Neural and Computational Viewpoints on Perceptual Decision-Making. Trends in Neurosciences 2018; 41: 838–52.

O’Dowd S, Schumacher J, Burn DJ, Bonanni L, Onofrj M, Thomas A, et al. Fluctuating cognition in the Lewy body dementias. Brain 2019; 142: 3338–50.

Onofrj M, Espay AJ, Bonanni L, Delli Pizzi S, Sensi SL. Hallucinations, somatic-functional disorders of PD-DLB as expressions of thalamic dysfunction. Mov Disord 2019; 34: 1100–11.

Peraza LR, Taylor J-P, Kaiser M. Divergent brain functional network alterations in dementia with Lewy bodies and Alzheimer’s disease. Neurobiology of Aging 2015; 36: 2458–67.

Philiastides MG, Sajda P. EEG-Informed fMRI Reveals Spatiotemporal Characteristics of Perceptual Decision Making. Journal of Neuroscience 2007; 27: 13082–91.

Power JD, Barnes KA, Snyder AZ, Schlaggar BL, Petersen SE. Spurious but systematic correlations in functional connectivity MRI networks arise from subject motion. NeuroImage 2012; 59: 2142–54.

Ratcliff R, McKoon G. The diffusion decision model: theory and data for two-choice decision tasks. Neural Comput 2008; 20: 873–922.

Ratcliff R, Smith PL, Brown SD, McKoon G. Diffusion Decision Model: Current Issues and History. Trends in Cognitive Sciences 2016; 20: 260–81.

Ratcliff R, Thapar A, McKoon G. Application of the Diffusion Model to Two-Choice Tasks for Adults 75−90 Years Old. Psychol Aging 2007; 22: 56–66.

Ratcliff R, Thapar A, McKoon G. Individual differences, aging, and IQ in two-choice tasks. Cognitive Psychology 2010; 60: 127–57.

Rissman J, Gazzaley A, D’Esposito M. Measuring functional connectivity during distinct stages of a cognitive task. NeuroImage 2004; 23: 752–63.

Roitman JD, Shadlen MN. Response of Neurons in the Lateral Intraparietal Area during a Combined Visual Discrimination Reaction Time Task. J Neurosci 2002; 22: 9475–89.

Roth M, Tym E, Mountjoy CQ, Huppert FA, Hendrie H, Verma S, et al. CAMDEX: a standardised instrument for the diagnosis of mental disorder in the elderly with special reference to the early detection of dementia. The British journal of psychiatry 1986; 149: 698–709.

Sauer J, Ffytche DH, Ballard C, Brown RG, Howard R. Differences between Alzheimer’s disease and dementia with Lewy bodies: an fMRI study of task-related brain activity. Brain 2006; 129: 1780–8.

Schumacher J, Peraza LR, Firbank M, Thomas AJ, Kaiser M, Gallagher P, et al. Dynamic functional connectivity changes in dementia with Lewy bodies and Alzheimer’s disease. NeuroImage: Clinical 2019a; 22: 101812.

Schumacher J, Peraza LR, Firbank M, Thomas AJ, Kaiser M, Gallagher P, et al. Dysfunctional brain dynamics and their origin in Lewy body dementia. Brain 2019b; 142: 1767–82.

Shadlen MN, Newsome WT. Neural Basis of a Perceptual Decision in the Parietal Cortex (Area LIP) of the Rhesus Monkey. Journal of Neurophysiology 2001; 86: 1916–36.

Shine JM, Muller AJ, O’Callaghan C, Hornberger M, Halliday GM, Lewis SJ. Abnormal connectivity between the default mode and the visual system underlies the manifestation of visual hallucinations in Parkinson’s disease: a task-based fMRI study. npj Parkinson’s Disease 2015; 1: 1–8.

Shine JM, O’Callaghan C, Halliday GM, Lewis SJG. Tricks of the mind: Visual hallucinations as disorders of attention. Progress in Neurobiology 2014: 1–8.

Spaniol J, Madden DJ, Voss A. A Diffusion Model Analysis of Adult Age Differences in Episodic and Semantic Long-Term Memory Retrieval. J Exp Psychol Learn Mem Cogn 2006; 32: 101–17.

Spiegelhalter DJ, Best NG, Carlin BP, Linde AVD. Bayesian measures of model complexity and fit. Journal of the Royal Statistical Society: Series B (Statistical Methodology) 2002; 64: 583–639.

Starns JJ, Ratcliff R. The effects of aging on the speed–accuracy compromise: Boundary optimality in the diffusion model. Psychology and Aging 2010; 25: 377–90.

Tiraboschi P, Salmon DP, Hansen LA, Hofstetter RC, Thal LJ, Corey-Bloom J. What best differentiates Lewy body from Alzheimer’s disease in early-stage dementia?= Brain 2006; 129: 729–35.

Tomassini A, Pollak TA, Edwards MJ, Bestmann S. Learning from the past and expecting the future in Parkinsonism: Dopaminergic influence on predictions about the timing of future events. Neuropsychologia 2019; 127: 9–18.

Voss A, Nagler M, Lerche V. Diffusion models in experimental psychology: A practical introduction. Experimental Psychology 2013; 60: 385–402.

Wagenmakers E-J. Methodological and empirical developments for the Ratcliff diffusion model of response times and accuracy. European Journal of Cognitive Psychology 2009; 21: 641–71.

Walker MP, Ayre GA, Cummings JL, Wesnes K, McKeith IG, O’Brien JT, et al. Quantifying fluctuation in dementia with Lewy bodies, Alzheimer’s disease, and vascular dementia. Neurology 2000a; 54: 1616–25.

Walker MP, Ayre GA, Cummings JL, Wesnes K, McKeith IG, O’Brien JT, et al. The Clinician Assessment of Fluctuation and the One Day Fluctuation Assessment Scale: Two methods to assess fluctuating confusion in dementia. Br J Psychiatry 2000b; 177: 252–6.

Walker MP, Ayre GA, Perry EK, Wesnes K, McKeith IG, Tovee M, et al. Quantification and Characterisation of Fluctuating Cognition in Dementia with Lewy Bodies and Alzheimer’s Disease. DEM 2000c; 11: 327–35.

Walpola IC, Muller AJ, Hall JM, Andrews-Hanna JR, Irish M, Lewis SJG, et al. Mind-wandering in Parkinson’s disease hallucinations reflects primary visual and default network coupling. CORTEX 2020; 125: 233–45.

Weil RS, Schrag AE, Warren JD, Crutch SJ, Lees AJ, Morris HR. Visual dysfunction in Parkinson’s disease. Brain 2016; 139: 2827–43.

Wiecki TV, Sofer I, Frank MJ. HDDM: Hierarchical Bayesian estimation of the Drift-Diffusion Model in Python. Front Neuroinform 2013; 7: 14.

Yau Y, Dadar M, Taylor M, Zeighami Y, Fellows LK, Cisek P, et al. Neural Correlates of Evidence and Urgency During Human Perceptual Decision-Making in Dynamically Changing Conditions. Cerebral Cortex 2020; 30: 5471–83.

Yeo BTT, Krienen FM, Sepulcre J, Sabuncu MR, Lashkari D, Hollinshead M, et al. The organization of the human cerebral cortex estimated by intrinsic functional connectivity. Journal of Neurophysiology 2011; 106: 1125–65.

Zarkali A, Adams RA, Psarras S, Leyland L-A, Rees G, Weil RS. Increased weighting on prior knowledge in Lewy body-associated visual hallucinations. Brain Communications 2019; 1: fcz007.

Zhang J, Rittman T, Nombela C, Fois A, Coyle-Gilchrist I, Barker RA, et al. Different decision deficits impair response inhibition in progressive supranuclear palsy and Parkinson’s disease. Brain 2016; 139: 161–73.

Zhang J, Rowe JB. Dissociable mechanisms of speed-accuracy tradeoff during visual perceptual learning are revealed by a hierarchical drift-diffusion model [Internet] Front Neurosci2014; 8[cited 2021 Jan 7] Available from: http://journal.frontiersin.org/article/10.3389/fnins.2014.00069/abstract.

