## Supplementary Material for "Impaired sensory evidence accumulation and network function in Lewy body dementia"

Comparison of DVARS and motion parameters across groups Page 2
*Supplementary Table 1.*

Comparison of number of no response trials across groups Page 3
*Supplementary Fig 1.*

Plot of posterior predictive against observed data Page 4
*Supplementary Fig. 2*

### 1. Comparison of DVARS and motion parameters

| | Controls | AD | LBD | *F* value | *p* value |
|---|---|---|---|---|---|
| **Mean xyz motion (mm)** | 0.14 (0.04) | 0.15 (0.06) | 0.18 (0.11) | 0.89 | 0.42 |
| **Max xyz motion (mm)** | 0.55 (0.22) | 0.82 (0.41) | 0.71 (0.47) | 1.73 | 0.19 |
| **Mean angular motion (°)** | 0.06 (0.01) | 0.11 (0.07) | 0.09 (0.08) | 2.51 | 0.09 |
| **Max angular motion (°)** | 0.46 (0.29) | 0.80 (0.59) | 0.59 (0.61) | 1.57 | 0.22 |
| **Mean DVARS** | 1.23 (0.11) | 1.26 (0.11) | 1.23 (0.087) | 0.65 | 0.52 |

Mean (standard deviation); Significance tests refer to between group one-way ANOVAs
AD = Alzheimer's disease; LBD = Lewy body disease.

**2. Comparison of number of no response trials across groups**

The amount of no response trials excluded from the analysis did not differ significantly across the groups [F(2, 35) = 1.97, $p$ = 0.154].

**Supplementary Figure 1.**

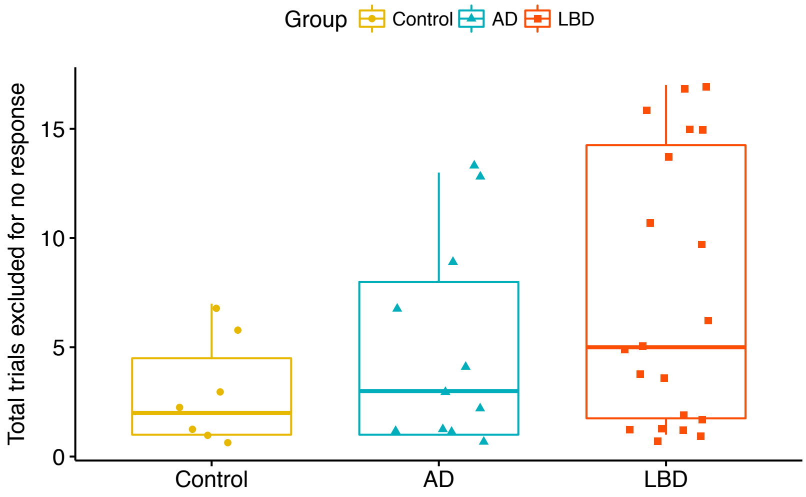

**3. Plot of posterior predictive against observed data**

Posterior predictive checks were conducted by averaging 500 simulations generated from the model's posterior to confirm it could reliably reproduce patterns in the observed data (Wiecki *et al.*, 2013).

**Supplementary Figure 2.**

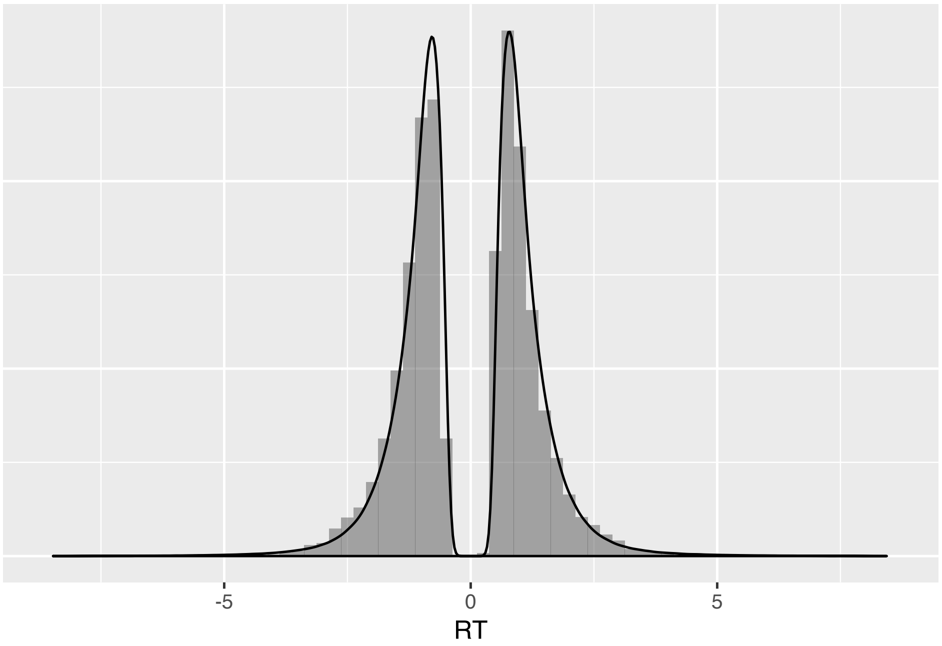


Histogram shows observed reaction time distribution (positive vs. negative results on the x-axis correspond to left vs. right responses). The black line plots the results from the posterior predictive simulation, i.e., data simulated based on using 500 parameter values from the posterior to simulate a different data set for each parameter value.

**References**

Wiecki TV, Sofer I, Frank MJ. HDDM: Hierarchical Bayesian estimation of the Drift-Diffusion Model in Python. Front Neuroinform 2013; 7: 14.
